# Real world hypertension treatment patterns analysis identifies putative gaps and risk-associated patterns

**DOI:** 10.1101/2020.10.28.20169623

**Authors:** Sricharan Bandhakavi, Jasmine M. McCammon, Sunil Karigowde, Zhipeng Liu, Xianglian Ni, Farbod Rahmanian

## Abstract

**Objective:** Although treatment patterns’ analyses at scale can provide insights into associated health outcomes, they remain relatively uncharacterized for most chronic diseases, including hypertension (HTN). To address this gap, we analyzed HTN treatment patterns among US health-insured patients.

**Materials and Methods:** New (n = 200,786) or all (n = 4.1 million) HTN patients were identified from 2015, 2016 enrollments in a nationwide administrative claims’ database and compared for HTN-specific treatment (Rx) choices, Rx count, and distinct rounds <u>o</u>f treatment <u>o</u>ptions (<u>ROTO</u>), respectively. Selected treatment patterns were risk-assessed using predictive modeling and/or literature-based recommendations.

**Results:** For 2016, ACE inhibitors/ARBs were most frequent choices in new HTN patients’ vs more diverse Rx-choices among all HTN patients. All HTN patients had ∼3-fold and ∼5-fold-higher prevalence of (same-year) polytherapy and Rx interventions, respectively.

New HTN patients with ≥2 rounds of treatment options (vs single/initial round) were associated with ∼5-fold or higher predicted HTN complications’ risk (p-value < 0.0001). All HTN patients with ≥3 rounds of treatment options (vs single/initial round) had 3.8-fold higher next-year HTN complications’ risk (p-value < 0.0001). Co-morbidities/persistence of ≥3 rounds of treatment options over 2 years further increased these odds and total medications/chronic disease score correlated with ROTO counts.

**Discussion:** ∼95% of new HTN patients in 2016 did not start treatment with current literature-recommended first option, Thiazides. Assuming “sticky” prescription patterns, opportunity exists to improve (current) initial HTN treatments. Additionally, ROTO counts can inform HTN complications’ risk/management thereof.

**Conclusion:** We highlight opportunities to improve initial HTN treatments and treatment patterns associated with higher risk among HTN patients.

## BACKGROUND AND SIGNIFICANCE

Among chronic diseases in the United States, hypertension (HTN) is one of the most prevalent [1]. According to the American Heart Association’s definition, there are more than 116 million adults in the United States with HTN; among adults ≥20 years, 49% of males and 42% of females are hypertensive in the US [1]. Given its high prevalence and its known association with a wide variety of other complications [2-4], HTN must be managed effectively in affected patients.

In addition to lifestyle interventions – *e.g*. through diet and exercise – there exist a variety of treatment (Rx) options for HTN management [5-7]. Benefits for each of the drug choices for HTN control has been demonstrated in various randomized control trial (RCT) studies done as part of initial drug applications for regulatory approval or subsequent to approval.

However, these RCT studies provide limited insights in the real world with respect to “ideal” option(s) along the patient’s treatment journey (based on their prior sequence of medications, number/classes of drugs per course, mix of comorbidities, age, gender etc.) and how various steps along the journey are associated with patient outcomes. For these insights, observational studies need to be done at scale to unravel potential relationships between treatment patterns/pathways and their health outcomes.

At least two major studies have been done to analyze HTN treatment patterns at scale in the United States (pre-2012 treatment patterns; [8]) and in China (pre-2015 treatment patterns; [9]). Interestingly, these studies identified significant differences between HTN treatment patterns in the US vs China. However, the identified patterns or their differences (across US vs China) were not subsequently analyzed for potential connections to health outcomes or concordance with literature/evidence-based recommendations. Thus, there is a need to extend these analyses further - to not only describe more current HTN treatment patterns but also elucidate their associated outcomes and concordance with current recommendations.

In this study, we characterized treatment patterns among newly progressed HTN vs all HTN patients enrolled for employer-based health insurance within the United States during 2015-2016. Identified patients were compared for initial vs subsequent/overall HTN-specific treatment (Rx) choices, frequency of monotherapy vs polytherapy, and number of distinct rounds <u>o</u>f treatment <u>o</u>ptions (ROTO) within calendar year, respectively. Selected treatment patterns were assessed further for future cost/risk using claims data, predictive modeling, and current literature-based recommendations. Based on these analyses, we highlight opportunities to improve initial HTN treatments and treatment patterns associated with higher risk among HTN patients.

## MATERIALS AND METHODS

### Identification of “newly progressed” vs “all” HTN patients

Newly progressed (or “new”) HTN patients were identified from an initial cohort of 20,251,628 patients who were enrolled in an Rx plan for 2015 and 2016 within a commercially licensed, (administrative) claims database [10 11]. Through the rest of this manuscript we refer to this source as the CCAE (<u>C</u>ommercial <u>C</u>laims <u>a</u>nd <u>E</u>ncounters) database. Within this cohort, patients were deemed to be “newly progressed” HTN patients if they did not pick-up any HTN-specific medications during 2015, but picked up HTN-specific medications anytime in 2016. All HTN patients in 2014, 2015, 2016 were also identified (without using the above filter of no HTN-specific medications in prior years), for subsequent comparative analyses. Patients taking HTN-specific medications were identified using the following ATC codes: C02, C03 except C03C, C04AB, C07, C08, C09 based on an earlier report [12]. **Figure 1A** summarizes this sequence of steps for identification of new versus all HTN patients in 2016.

**Figure 1.**
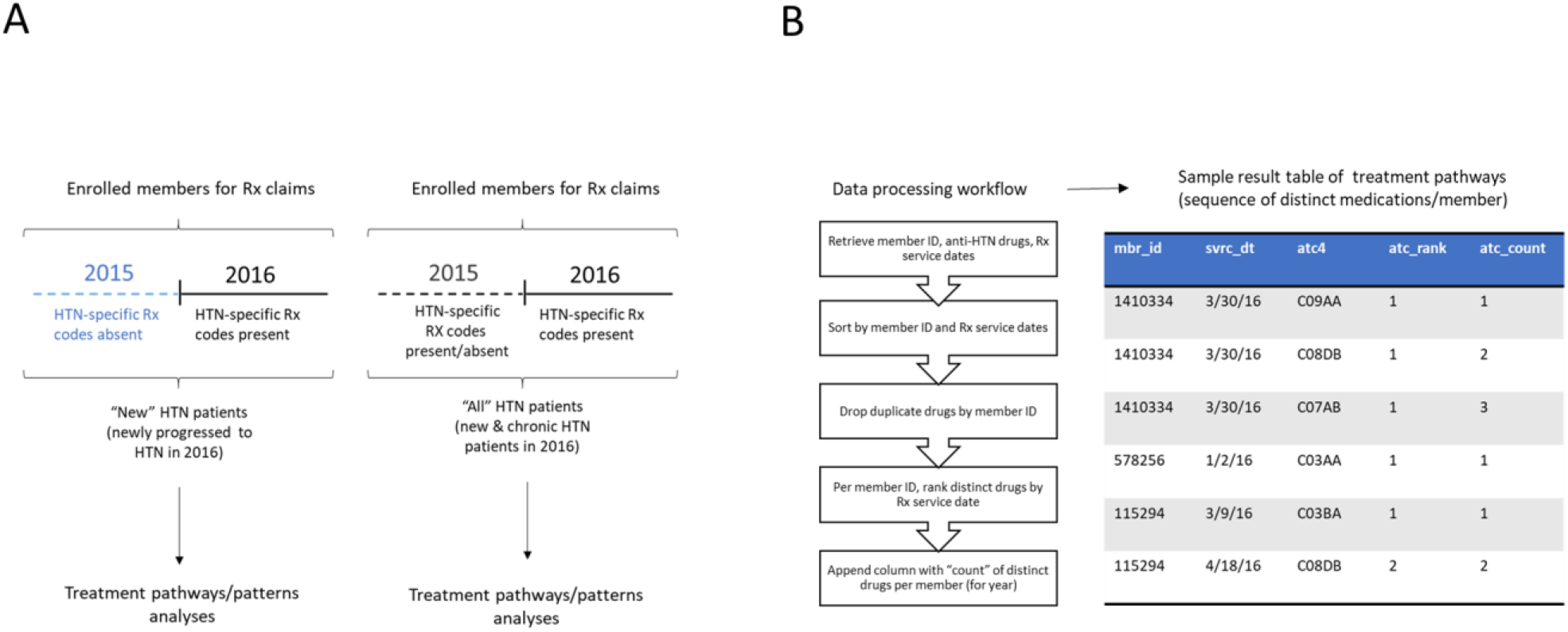
Patient cohort selection strategy and data processing workflow. **A**. Patients newly progressed to HTN in 2016 vs all HTN patients in 2016 were selected from those enrolled through 2015 and 2016. New HTN patients were identified based on absence of HTN-specific Rx codes in 2015 (but not in 2016). HTN-specific Rx codes used were: C02, C03 except C03C, C04AB, C07, C08, C09. **B**. Data processing workflow and resulting sample treatment pathways representation; *mbr_id* = member ID, *svrc_dt* = date of Rx pick-up, *atc4* = drug class based on 5-level anatomical therapeutic chemical (ATC) classification system, *atc_rank* = initial vs subsequent round(s) of treatment options (ROTO), *atc_count* = count of distinct drugs per patient for calendar year.

Using preceding filtering criteria, 200,786/20,251,628 members (∼1% of total enrolled) were identified as “newly progressed” or “new HTN” patients and 4,113,781/20,251,628 members (∼20% of total enrolled) were identified as “all HTN” patients in 2016. Thus, ∼4.9% (200,786/4,113,781) of all HTN patients were “newly progressed” to HTN indicating that the vast majority (>95%) of “all HTN” patients may be considered as having “chronic”/pre-existing HTN.

### Treatment pathways “dataframe” construction

A tabular representation of data (“dataframe”) of HTN-specific treatment pathways i.e., sequence of distinct HTN-specific medications for “new HTN” vs “all HTN” members was constructed using the workflow outlined in **Figure 1B** *(left panel)* and a sample abbreviated “dataframe” is shown in **Figure 1B** *(right panel)*. The treatment pathways “dataframe” was further processed extensively using Python 3.6.1/SQL scripts for the results in this report. For selected analyses, co-morbidity measures - CCI (<u>C</u>harlson <u>C</u>omorbidity Index) and CDS (<u>C</u>hronic <u>D</u>isease <u>S</u>core) values for 2015 were calculated based on 2015 medical claims data [13] and 2015 pharmacy claims data [14 15], respectively. Odds-ratios were calculated using contingency table/chi-squared analysis using python scripts and an online odds-ratio calculator (https://www.medcalc.org/calc/odds_ratio.php).

### HTN complications predictive model

Predictive models were generated in-house using claims data from the CCAE database to predict for those with HTN, who are likely to develop HTN complications of increasing severity (from stage 1 being least severe to stage 3 being most severe) in the next 12 months. HTN complications were grouped as stage 1/stage 2/stage 3 complications based on underlying medical claim codes. The underlying medical codes used for grouping complications based on severity and overall modeling strategy details are described in an accompanying manuscript [16]. The most relevant details for this report are outlined below.

#### Patient cohort

Patient cohort consisted of those enrolled in 2015 and 2016 within the CCAE database and who had at least 2 HTN-specific Rx claims and 2 HTN-specific medical claims (n = ∼1.6 million). Among them, patients with vs without HTN complications in 2016 grouped by stage were identified based on underlying ICD-10 diagnosis codes and labeled as 1 vs 0 (referred to as “Y/outcome variable”) in predictive modeling.

#### Features

A total of 96 features (collectively also referred to as “X matrix”) were generated using SQL/python for the above patient cohort using 2015 claims data. Features spanned across demographics (age, sex), weight of comorbidities (CCI), prescriptions (for HTN, comorbidities, and interfering medications), health service utilization (costs and medical visits/admits), relevant procedure and diagnosis codes.

#### Predictive modeling workflow

Features (“X matrix”) and outcome variable (“Y”) from above were combined to generate a dataset of ∼1.6 million total records. This dataset was split 75:25 into train: test samples using stratified sampling. An optimized, hyper-parameter tuned, logistic regression algorithm was trained on 75% of the data (train sample), performance tested on the remaining 25% of data (test sample), and further validated using an independent dataset of commercially insured patients from a distinct payer. Model performance against test sample (AUROC, sensitivity, and specificity) were as follows; stage 1 HTN complications: (0.78, 0.62, 0.81), stage 2 HTN complications: (0.87, 0.72, 0.89), and stage 3 HTN complications: (0.84, 0.69, 0.85), respectively.

## RESULTS

This study was performed in three stages. In the first stage, we focused on “new HTN” patients in 2016 (i.e., newly progressed in 2016) and their treatment pathways/patterns. Within these findings, selected observations were further evaluated against most recent literature/evidence-based guidelines as described in the discussion section. In the next stage, we compared treatment pathways/patterns at an aggregate level of “new HTN” patients with those of “all HTN” patients. Finally, based on above comparisons, we performed a risk/cost characterization of selected patient sub-groups. Results from each of these analyses are presented below.

**Figure 1** outlines the patient cohort selection strategy and data processing workflow for new HTN patients’ vs all HTN patients in 2016. As shown in **Figure 1A** *(left panel)*, new HTN patients were identified based on their enrolment through 2015 and 2016, and absence of any HTN-specific medication pick-ups in 2015 but not in 2016. By contrast, all HTN patients may have had HTN-specific Rx claims in 2015 also – i.e., they included new HTN patients (<5%) and chronic/previously hypertensive patients (>95%). Patient claims for each group (new HTN or all HTN) were processed further using the workflow shown in **Figure 1B** *(left panel)* resulting in a “dataframe” (*i.e*., tabular representation) of the sequence of distinct medications for each patient.

A fictitious limited “example” of the “dataframe” is shown in **Figure 1B** *(right panel)* wherein member IDs, Rx service dates and (non-hypertensive) drug classes by ATC4 codes are listed, along with two additional features that are directly relevant to “treatment pathways” analyses per member. These are: (1) *atc_rank* – refers to total number of distinct rounds of treatment Rx interventions/changes; we also refer to this as rounds <u>o</u>f treatment <u>o</u>ptions (ROTO) within this report, and (2) *atc_count* – which refers to total number of distinct anti-HTN medications taken in 2016 calendar year. For example, the first member (*mbr_id*: 1410334) had a single set of medications throughout the year (single round <u>o</u>f treatment <u>o</u>ptions/SROTO with three distinct medications). The next member *(mbr_id*: 572856) also is a SROTO member but stayed on monotherapy through the year. And finally, the last member (*mbr_id*: 115294) had <u>m</u>ultiple rounds <u>o</u>f treatment <u>o</u>ptions (MROTO) with two distinct medications over two ROTOs (ATC code C03BA for initial ROTO and ATC code C08DB as an Rx change/addition in subsequent ROTO). The new HTN patients’ treatment pathways “dataframe” was analyzed initially next.

**Figure 2** shows the frequency of various Rx options used in the new HTN patients and how these changed by each treatment round/subsequent intervention. As shown in **Figure 2A** (*left panel*), the most common drugs used for new HTN patients were ACE inhibitors and <u>A</u>ngiotensin <u>R</u>eceptor <u>B</u>lockers (ARBs) which target the RAAS/renin <u>a</u>ldosterone <u>a</u>ngiotensin system [17]. **Figure 2A** (*right panel*) shows that ACE inhibitors were the first Rx choice for ∼68% of new HTN patients. By contrast, <5% of the new HTN patients started with thiazides.

**Figure 2.**
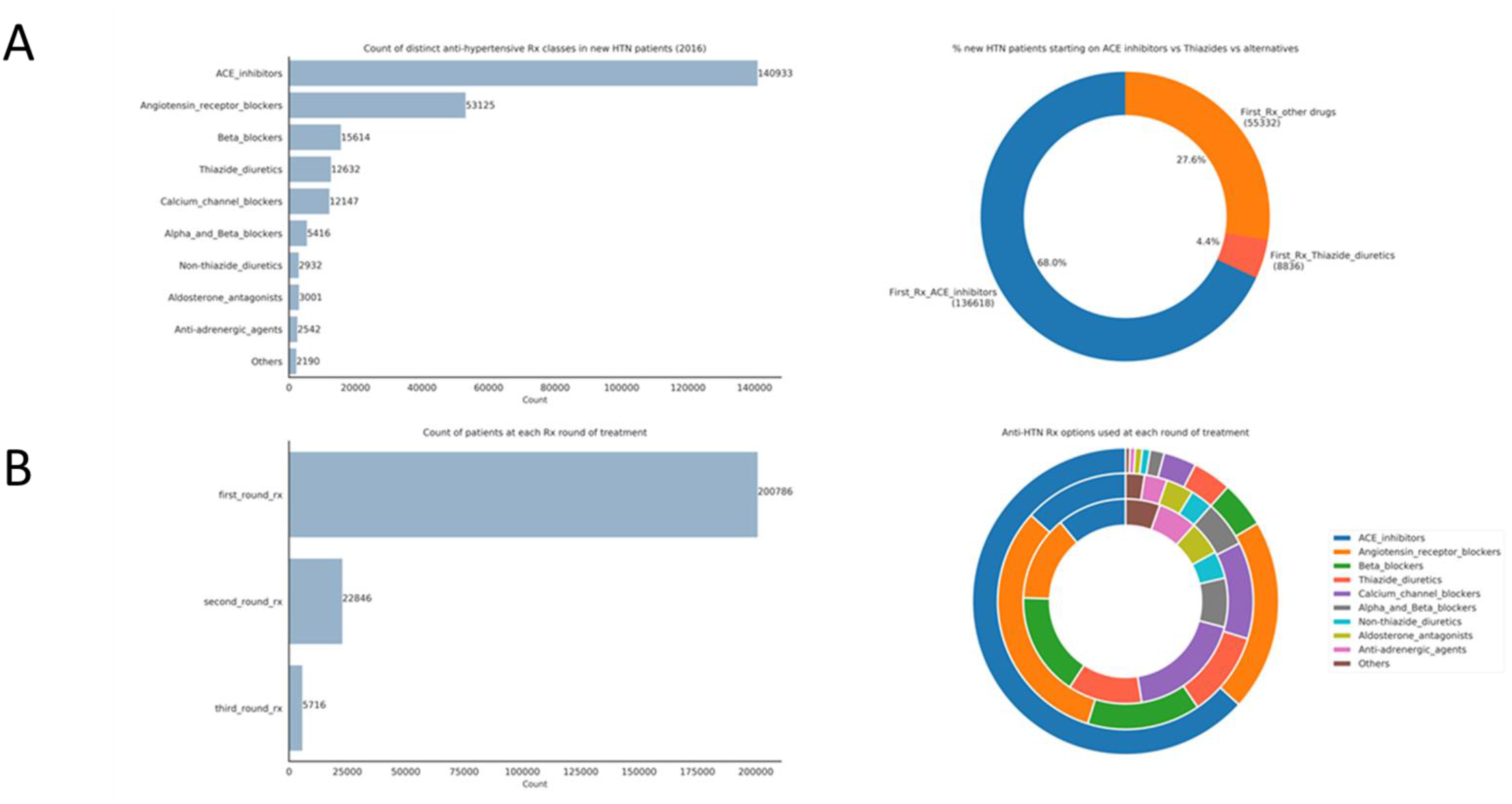
Treatment patterns among new HTN patients in 2016. **A**. Count of distinct medication classes for new HTN patients in 2016 and percentage of patients initiating therapy on selected anti-hypertensive medication classes. **B**. Count of patients at each round of treatment and frequency of Rx options used within. Outer circle represents relative frequencies in “*first_round_rx*” options, middle circle represents relative frequencies in “*second_round_rx*” options, and innermost circle represents relative frequencies in “*third_round_rx*” options, respectively.

Next, we further analyzed the number of patients needing subsequent changes to their initial HTN therapies and their Rx choices in each change/round of treatment option(s). As shown in **Figure 2B** (*left panel*), about ∼11% of new HTN patients (22,000 out of 200,786) had a subsequent Rx intervention *i.e*., Rx change/addition (*second_round_rx*) and 5,716 of these patients (∼3% of all new HTN patients, *third_round_rx*) needed two subsequent Rx interventions since the initial *first_round_rx*. **Figure 2B** (*right panel*) shows the relative frequency of anti-hypertensive medications in each of these treatment “rounds” across all the patients. As indicated, ACE inhibitors and ARBs were the predominant first “round” Rx options. The second “round” of anti-HTN options showed a much lower use of ACE inhibitors and ARBs, although ARBs remained prominent. In the third “round”, anti-HTN drug choices were more diverse, with no clear favorite among several prominent options including ACE inhibitors, ARBs, <u>B</u>eta <u>B</u>lockers (BBs), and <u>C</u>alcium <u>C</u>hannel <u>B</u>lockers (CCBs).

In the next stage of our analysis (see **Figure 3**), we compared treatment patterns – across types/number of drugs and number of Rx interventions/rounds <u>o</u>f treatment <u>o</u>ptions (ROTOs) - for new HTN vs all HTN patients in 2016 to unravel potential differences of further interest. As shown in **Figure 3** (*left panel*), ACE inhibitors and ARBs are most common Rx options among new HTN patients. By contrast, all HTN patients have a more diverse selection of Rx choices – with up to 5 different drug classes that are roughly equivalently popular. Compared to new HTN patients, all HTN patients had ∼5-fold higher prevalence of polytherapy (the vast majority of which are on dual therapy; **Figure 3**, *middle panel*) and ∼3-fold higher prevalence of multiple rounds of treatment options (MROTO vs SROTO; **Figure 3**, *right panel*), respectively. Thus, as HTN progresses to a more chronic condition, therapeutic patterns/complexity increases as evidenced by more diverse drugs choices (that are reminiscent of the third round treatments in new HTN patients; **Figure 2B**, *right panel*), along with higher prevalence of polytherapy and multiple Rx interventions/ROTOs.

**Figure 3.**
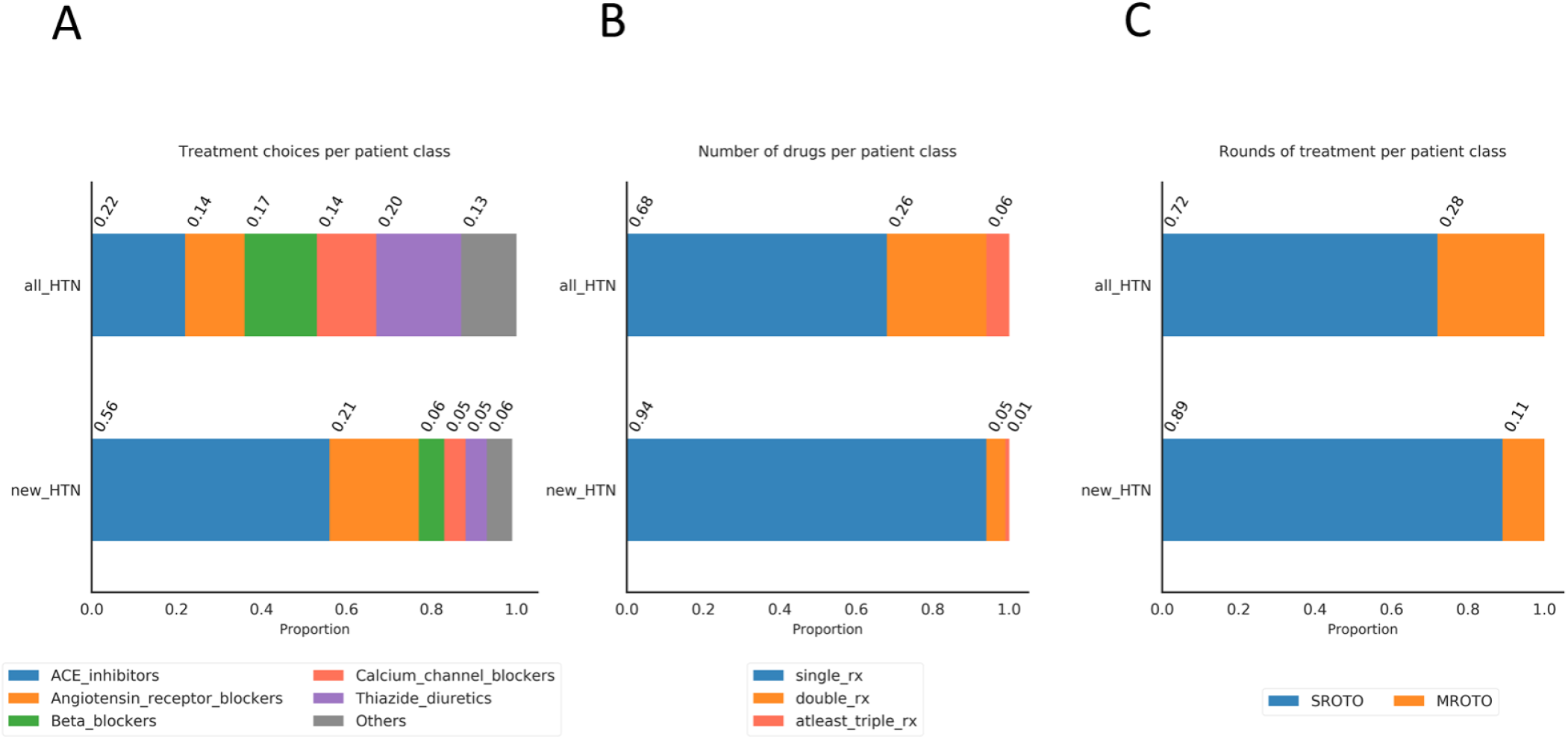
Comparison of treatment choices or treatment patterns in all HTN vs new HTN patients. **A**. Percentage of patients on each treatment choice over calendar year. **B**. Percentage of patients on single vs multiple (i.e., two distinct drug classes or at least three distinct drug classes) anti-HTN drugs over calendar year. **C**. Percentage of patients on single vs <u>m</u>ultiple rounds <u>o</u>f treatment <u>o</u>ptions (SROTO/MROTO).

We investigated further, the potential relationship between (anti-hypertensive) drug counts/year or Rx interventions/ROTO class on current and future risk in HTN patients (**Figure 4**). For this investigation, we leveraged CCAE claims-based models that predicted probability of HTN complications of increasing severity from stage 1 to stage 2 and stage 3 in 2016 (see also Materials and Methods) based on 2015 claims. Specifically, we visualized the probability distributions for each HTN complication stage returned by these models as box-plots against drug counts or ROTO class for new HTN patients (**Figure 4A**) and ROTO class for all HTN patients (**Figure 4B**).

**Figure 4.**
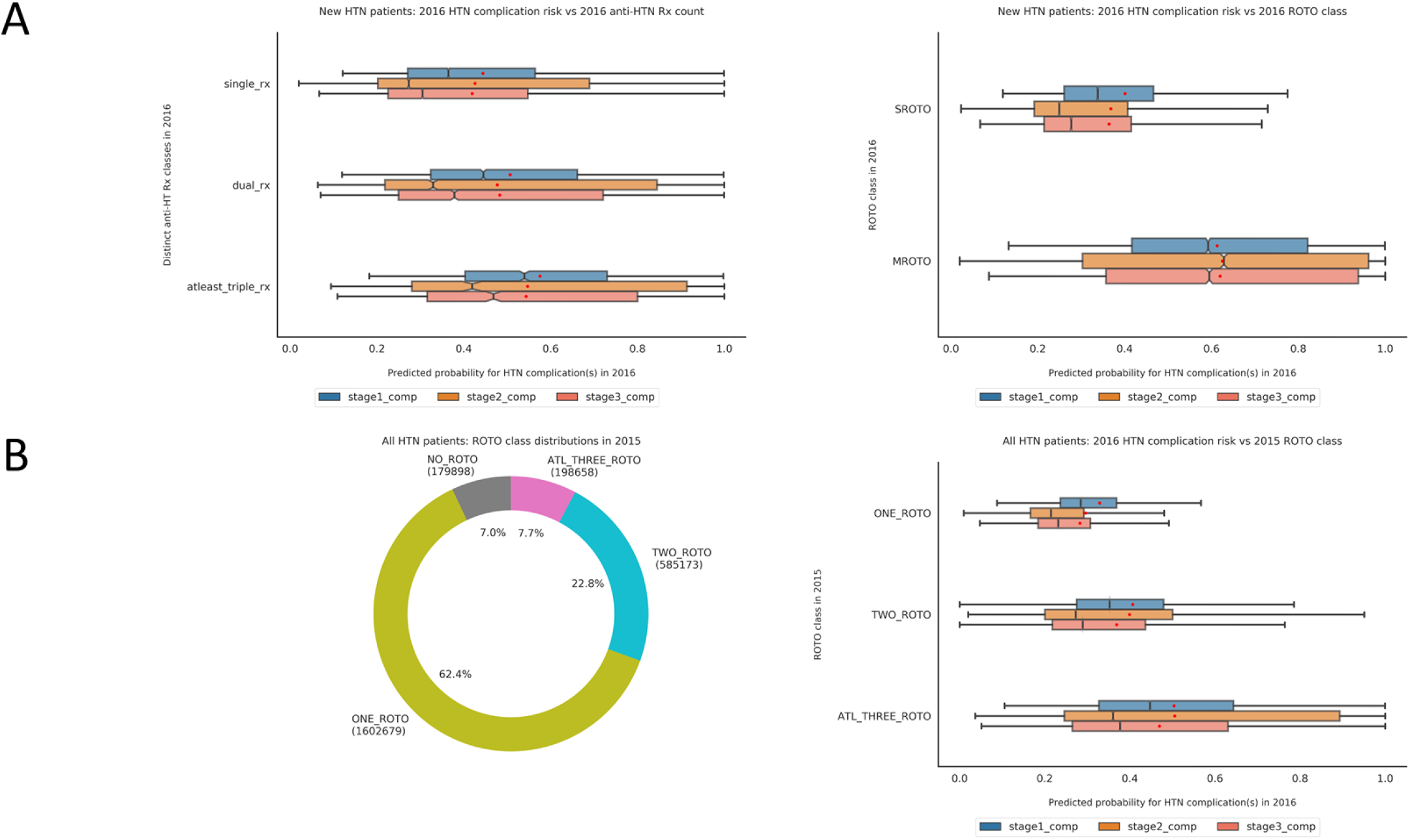
Predicted HTN complication risk for new HTN and all HTN patients based on selected treatment patterns. Notched box-plots ae used to visualized probability distributions wherein the “notch” displays the 95% confidence interval around median, red dot indicates the average and outliers were not visualized (although included for generation of statistics on box-plots). **A**. Same year (2016) HTN complication risk in new HTN patients’ vs number of distinct anti-HTN drug classes taken in 2016 *(left panel)* or number of single/<u>m</u>ultiple rounds <u>o</u>f anti-HTN treatment <u>o</u>ptions (SROTO/MROTO; *right panel*) in 2016. **B**. Distribution of all HTN patients by rounds of treatment in 2015 (*left panel*) and their predicted 2016 HTN complications risk (*right panel*); NO_ROTO = <u>no r</u>ounds <u>o</u>f treatment <u>o</u>ptions in 2015, ONE_ROTO = <u>one r</u>ound <u>o</u>f treatment <u>o</u>ptions (i.e., no Rx changes/additions in same year) in 2015, TWO_ROTO = two rounds <u>o</u>f treatment <u>o</u>ptions in 2015, and ATL_THREE_ROTO = <u>at l</u>east <u>three r</u>ounds <u>o</u>f treatment <u>o</u>ptions in 2015.

As shown in **Figure 4A** (*left panel*), there is a modest rightward shift in the predicted probabilities for stage 1, stage 2, and stage 3 HTN complications in new HTN patients in 2016 as their number of drugs/year increased from one (*single_rx*), to two (*dual_rx*), and at least three (*atleast_triple_rx*). Relative to the comparison with number of drugs, there is an even more dramatic shift in predicted probabilities of HTN complications for MROTO vs SROTO patients (**Figure 4A**, *right panel*). The “notched” medians within the box-plots represent the 95% CI value of the median estimated by bootstrapping; non-overlapping notches/medians indicate statistically significant differences in the median probabilities across groups being compared.

Given the more dramatic association seen with ROTO class (vs number of drugs) for predicted HTN risk, we further analyzed the association between ROTO class and predicted future HTN risk in all HTN patients. **Figure 4B** shows the distribution/prevalence of members with each ROTO class among all HTN patients in 2015 and their predicted (future) probabilities for HTN complications in 2016. To better capture the potential relationship between these features (past year ROTO class vs future HTN complications risk), we discretized all HTN patients into 4 categorical bins – NO_ROTO in 2015 (<u>no r</u>ounds <u>o</u>f treatment <u>o</u>ptions in 2015), ONE_ROTO (<u>one r</u>ound <u>o</u>f treatment <u>o</u>ptions in 2015 *i.e*., had no subsequent changes/additions to their HTN medications within same year), TWO_ROTO (<u>two r</u>ounds <u>o</u>f treatment <u>o</u>ptions i.e., had one subsequent Rx intervention as a change/addition to initial HTN treatment/s in 2015) and ATL_THREE_ROTO (<u>at l</u>east <u>three</u> rounds <u>o</u>f treatment <u>o</u>ptions i.e., had at least two subsequent Rx interventions as changes/additions since initial HTN treatment/s in 2015).

As shown in **Figure 4B** (*left panel*), the largest % of all HTN patients in 2015 belonged to the ONE_ROTO class (62.5%), followed by the TWO_ROTO class (22.8%). Approximately 7% of the patients had no Rx treatments (NO_ROTO class members) or had at least three ROTOs in 2015 (ATL_THREE_ROTO class members). **Figure 4B** (*right panel*) shows the relationship of predicted probabilities for 2016 HTN complications for all HTN patients who belonged to one of the four ROTO classes in 2015. 2015 ATL_THREE_ROTO class members have the most “right-shifted” probability distributions of HTN complications in 2016 across all stages (stage 1, stage 2, and stage 3) indicating the highest predicted future risk among all ROTO classes.

To quantify the increased predicted HTN complications risk by ROTO class, we generated predicted odds of HTN complications based on the probability cut-off values/thresholds used by our HTN complications’ predictive models. For the performance metrics described in the materials and methods sections, the HTN complications models used a probability threshold of 0.47 for stage 1 HTN complications, 0.431 for stage 2 HTN complications, and 0.498 for stage 3 HTN complications; all patients who had a model-generated probability score higher than the preceding probability thresholds were “predicted” to develop stage 1/2/3 HTN complications in 2016, respectively. Using these probability thresholds, the *<u>relative odds</u>* of same-year (2016) predicted HTN complications for MROTO (vs SROTO) new HTN patients were as follows: [*<u>6.19</u>*, 95% CI: 5.93 - 6.46, p-value < 0.001] for stage 1 HTN complications, [*<u>4.65</u>*, 95% CI: 4.46 - 4.84, p-value < 0.001] for stage 2 HTN complications, and [*<u>5.97</u>*, 95% CI: 5.72 - 6.23, p-value < 0.001] for stage 3 HTN complications. The *<u>relative odds</u>* of next-year (2016) predicted HTN complications based on 2015 ROTO class for ATL_THREE_ROTO (vs ONE_ROTO) all HTN patients were as follows: [*<u>10.31</u>*, 95% CI: 10.21 – 10.42, p-value < 0.001] for stage 1 HTN complications, [*<u>5.77</u>*, 95% CI: 5.72 – 5.83, p-value < 0.001] for stage 2 HTN complications, and [*<u>8.5</u>*, 95% CI: 8.41 – 8.59, p-value < 0.001] for stage 3 HTN complications. Thus, increasing ROTO counts significantly increased same year risk predictions for HTN complications in new HTN patients and future/next year risk predictions for HTN complications in all HTN patients.

Although preceding results implicate ROTO counts as a potential risk factor associated with HTN complications, they are based on the probability thresholds of a predictive model and not actual counts of HTN complications. Hence, as an independent validation, we next used actual counts of HTN complications (regardless of stage of complication) to evaluate the strength of the potential relationship between ROTO counts and future risk. Specifically, we sought to determine if ROTO counts in prior year affected overall (future) total costs as well or mainly drove the (future) risk of HTN complications. Finally, we investigated the potential role of co-morbidities (using <u>C</u>harlson <u>C</u>omorbidity Index/CCI [13]) and/or co-medications’ complexity (using <u>C</u>hronic <u>D</u>isease <u>S</u>core/CDS [14 15]) in the relationship between ROTO class and future HTN complications. The results of these analyses are shown in **Figures 5A, 5B**.

**Figure 5.**
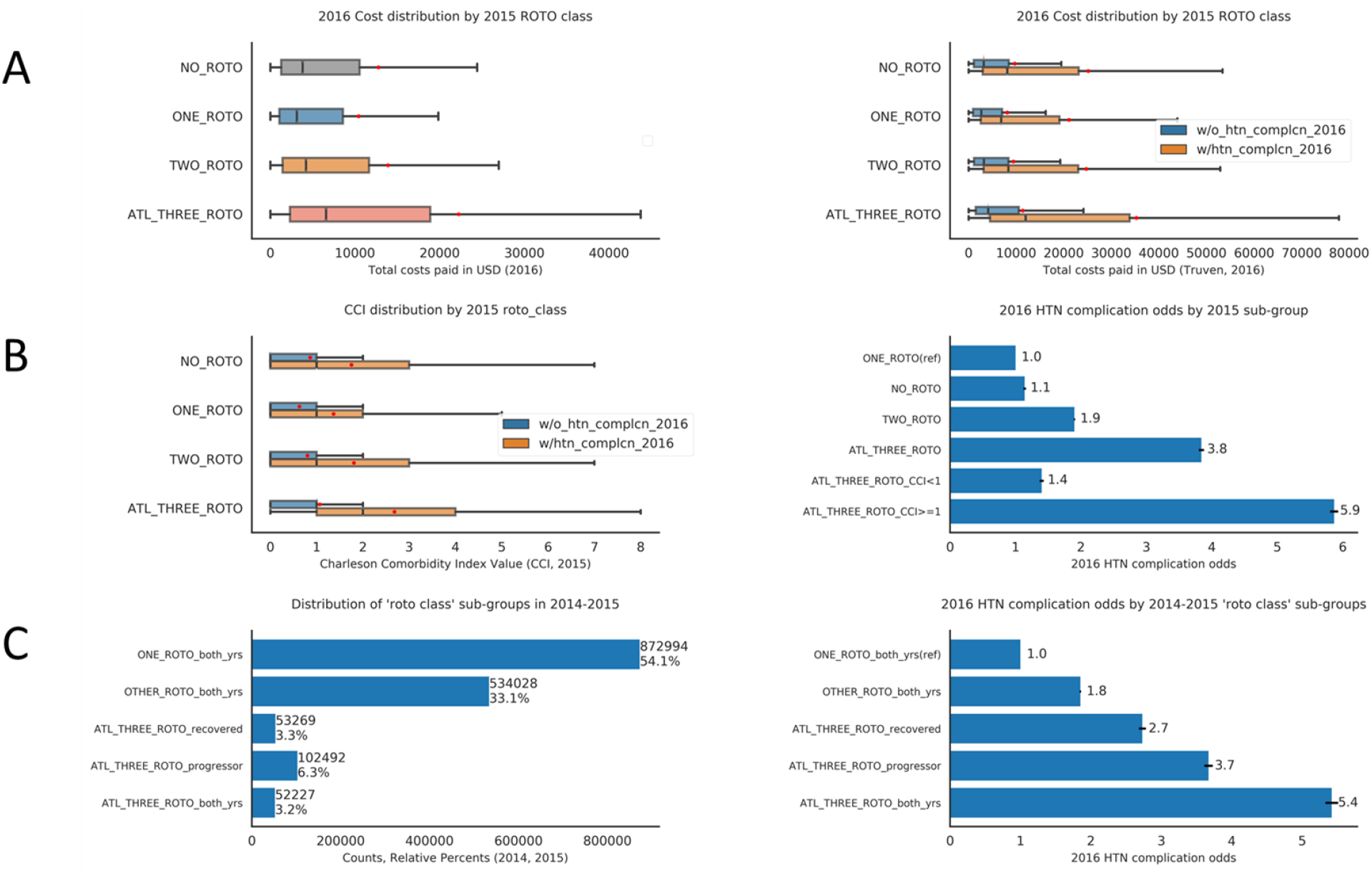
Patient sub-group analyses and correlations with HTN complications. **A**. 2016 cost distributions by 2015 ROTO class overall (*left panel*) vs after ROTO class stratification by observed HTN complications in 2016 (*right panel*). Number of patients who developed HTN complications vs not are indicated within parentheses in right panel and notched box-plots used to visualize the distributions. **B**. Same-year (2015) CCI distributions in 2015 ROTO groups stratified by observed HTN complications in 2016 (*left panel*) and odds of HTN-complications in 2016 for selected 2015 patient sub-groups (*right panel*); error bars for odds ratios indicate 95% confidence intervals and CCI = <u>C</u>harlson <u>C</u>omorbidity Index value. Notched box-plots are used to visualize distributions. **C**. Distribution of ROTO classes over 2014-2015 via notched-box plot analyses (*left panel*) and association with HTN complications in 2016 (*right panel*); error bars for odds ratios indicate 95% confidence intervals.

As shown in **Figure 5A** *(left panel)*, ROTO class in 2015 did not substantially increase next-year total costs except in the case of ATL_THREE_ROTO patients. Regardless of ROTO class, patients with HTN complications in 2016 had 2-3 fold higher median total costs in 2016 (**Figure 5A**, *right panel*). Interestingly, in the absence of future HTN complications, the total cost of ATL_THREE_ROTO patients is roughly comparable to that of other ROTO classes without HTN complications (**Figure 5B**, *right panel*); thus, the increased future costs in ATL_THREE_ROTO patients are mostly driven by future HTN complications.

There is a modest increase in median number of co-morbidities (measured by 2015 CCI scores) as 2015 ROTO count increases; this correlation is even stronger for 2015 chronic disease score – a measure of co-medication burden across 27 disease conditions (see **Supplementary Figures 1A, 1B**). Across all 2015 ROTO classes, 2015 CCI score was often higher for those who developed HTN complications in next year (**Figure 5B**, *left panel*). This is most striking in the case of 2015 ATL_THREE_ROTO patients wherein ∼75% of ATL_THREE_ROTO patients who developed HTN complications in the next year had at least 1 co-morbidity (CCI value of ≥1); by contrast, ∼75% of ATL_THREE_ROTO patients who did NOT develop HTN complications in the next year, had no co-morbidities (**Figure 5B**, *left panel*). Thus, CCI values can “separate” majority of ATL_THREE_ROTO patients who developed HTN complication next year vs not. CDS scores do not show a similarly pronounced ability to “separate” ATL_THREE_ROTO patients who developed HTN complications next year; instead CDS scores are positively correlated with ROTO count (**Supplementary Figures 1B, 1C**).

Next, we used odds ratios to quantify the relationship between 2015 ROTO class and/or CCI with HTN complications in 2016. As shown in **Figure 5B** *(right panel)*, actual odds of (any stage) HTN complications in 2016 were highest (3.8-fold vs ONE_ROTO, p-value <0.0001) in ATL_THREE_ROTO patients among all 2015 ROTO classes. Presence of any co-morbidities (CCI ≥1) increased these odds further (5.9-fold vs ONE_ROTO, p-value < 0.0001) and absence of any co-morbidities reduced the odds of future HTN complications in ATL_THREE_ROTO patients substantially even though they remained statistically significant (1.4-fold vs ONE_ROTO, p-value < 0.0001). Thus, ATL_THREE_ROTO status in all HTN patients drives future HTN complications risk independently *and* in combination with pre-existing co-morbidities.

As an additional measure of evaluating the putative association between ATL_THREE_ROTO status and future HTN complications risk, we determined the relative persistence of ATL_THREE_ROTO status over a 2-year observation period (2014-2015) among all HTN patients and its associated odds of HTN complications in 2016. As shown in **Figure 5C** (*left panel*), the majority (*ONE_ROTO_both_yrs*; ∼54%) maintained ONE_ROTO status (i.e., did not need any subsequent Rx interventions in each calendar year) over the 2014-2015 period. The next largest group of patients did not achieve ATL_THREE_ROTO status in either of the two years (*OTHER_ROTO_both_yrs*; ∼33%). The remaining patients achieved ATL_THREE_ROTO status in either 2014 but not 2015 (*ATL_THREE_ROTO_recovered*; 3.3%) or 2015 only (*ATL_THREE_ROTO_progressor*; 6.3%) or both 2014 and 2015 (*ATL_THREE_ROTO_both_yrs*; 3.2%). Importantly, patients with persistent (*ATL_THREE_ROTO_both_yrs*) or the most recent ATL_THREE_ROTO status (*ATL_THREE_ROTO_progressor*) had highest odds of future/next-year HTN complications whereas those who recovered one year ago (*ATL_THREE_ROTO_recovered*) had lower, but still significant risk (**Figure 5C**, *right panel*). Thus, the reduction of ATL_THREE_ROTO status is not only possible but beneficial, and ATL_THREE_ROTO status informs future risk over a two-year window, at least.

## DISCUSSION

In this study, we have characterized treatment patterns among newly progressed HTN patients’ vs all HTN patients enrolled for employer-based health insurance within the United States. Selected treatment patterns were assessed further for future cost/risk using claims data, predictive modeling, and as discussed further in this section, current literature-based recommendations. Taken together, our analyses have identified opportunities to improve initial treatments for new HTN patients and highlight treatment patterns associated with current/future HTN complications’ risk. These notions are evaluated further below.

An important question for HTN patients/providers has been to determine the optimal first Rx choice for newly progressed HTN patients. However, current guidelines provide only limited guidance on which Rx option to initiate therapy with for new HTN patients [5-7]. Towards addressing this current guidance “limitation”, two recent meta-analyses [18 19] are particularly relevant and both meta-analyses report that initiating therapy for HTN with low-dose thiazides is beneficial. Among these, the LEGEND-HTN study [18] stands out as the most comprehensive analysis to date; it included pair-wise comparisons for efficacy & safety across 4.9 million patients starting on monotherapy with one of five different first-line drug classes having a median follow-up duration of > 2 years. Patients starting with thiazides/thiazide-like diuretics had a significantly lower risk for 43/46 total safety outcomes and 3/3 primary effectiveness outcomes of the study: acute myocardial infarction, hospitalization for heart failure, and stroke - as compared with those starting & staying on one of two drug choices: ACE inhibitors or non-dihydropyridine CCBs.

Thiazides – mainly when used at high doses – have been connected to statistically significant increases in blood glucose levels although the clinical significance of this modest increase has remained somewhat unclear [20 21]. However, this effect on blood glucose levels has limited their use (relative to ACE inhibitors) especially in patients susceptible to/diagnosed with Diabetes [20]; this Rx preference likely also extends to initial Rx choice for newly progressed HTN patients. Notably, the LEGEND-HTN study indicated the superiority of starting therapy with Thiazides over ACE inhibitors *<u>after</u>* adjusting for Diabetes and several other co-variates (patients were matched across 43 medical conditions, 18 medication classes, age, gender). An important implication of the LEGEND-HTN study is that 1·3 cardiovascular events could be avoided for every 1000 patients who initiated treatment with a thiazide or thiazide-like diuretic instead of an ACE inhibitor.

Although the above studies came out in 2018 and 2019 (i.e., are more recent than the 2015-2016 period for which we analyzed treatment pathways of new HTN patients), treatment patterns can remain “sticky” over many years, unless revised by guidelines. Given the absence of guideline revisions since 2017 and the safety implications of the LEGEND-HTN study, we investigated the percentage of new HTN patients who initiated therapy with ACE inhibitors vs Thiazide diuretics in our analysis. Strikingly, less than 5% of new HTN patients in our study started therapy with the Thiazide diuretics (from the >90% who started on monotherapy). The vast majority (68%) of new HTN patients started with ACE inhibitors – a drug choice shown to be less favorable than Thiazides in the LEGEND-HTN study [18]. Indeed, the LEGEND-HTN study also reported that < 18% of new HTN patients who initiated with monotherapy started on Thiazides or Thiazide-like diuretics and ∼50% started on ACE inhibitors; however, these percentages were reported over the entire patient cohort accumulated across 1996 – 2018 in multiple databases. By contributing this relatively recent prevalence information (for use of thiazides vs ACE inhibitors as initial starting anti-HTN therapy in 2016), and assuming stickiness of prescription patterns since then, our report highlights a potential opportunity to improve patient outcomes – for new HTN patients – by increasing the use of Thiazides/Thiazide-like diuretics over ACE inhibitors in the United States.

Additionally, we have analyzed treatment patterns among new HTN and all HTN patients (in our 2015 – 2016 study cohort) that might inform current or future risk of HTN complications. For this analysis, we leveraged a combination of predictive modeling and as an independent validation, used actual counts/events of HTN complications in 2016. This identified significant association for number of rounds <u>o</u>f treatment <u>o</u>ptions (ROTO) with same-year HTN complications in new HTN patients and same-year & future HTN complications in all HTN patients. Among all HTN patients, ∼8% of all HTN patients had <u>at l</u>east <u>three r</u>ounds <u>o</u>f treatment <u>o</u>ptions (ATL_THREE_ROTO) in same calendar year and these were associated with 3.8-fold or higher odds of future HTN complications. They are also associated with elevated same-year risk (see **Supplementary Figure 2**). Thus, patients with elevated ROTO counts – especially ATL_THREE_ROTO class - likely represent a sub-group in whom HTN is challenging to control requiring multiple Rx adjustments/changes and who progress to more serious outcomes in the near future.

Patients who reduced their ROTO count from ATL_THREE_ROTO status over a two-year period reduced their future HTN complications risk whereas those who did not, increased their risk further. This information can be exploited by clinical/risk management programs to identify high-risk patients over a two-year window who might benefit from targeted care-management interventions (e.g. via monitoring treatment efficacy/options, medication adherence, lifestyle interventions, and possibly other ideas not explored here). For such interventions, high risk ATL_THREE_ROTO patients may be initially targeted based on their CCI score/existing co-morbidities or persistence of ATL_THREE_ROTO status over multiple years or both, and further prioritized - to potentially increase the likelihood of successful interventions - by focusing initially on those patients having relatively low CDS scores (i.e., less complicated treatment regimens). Outside of targeting patients by ROTO status, it may also be used to monitor efficacy of HTN care programs overall – wherein high risk patients are identified via predictive modeling *en masse* [16] and patients monitored for their ROTO status and/or reduction of ROTO count (if initially belonging to ATL_THREE_ROTO class). Further work is needed to validate these notions in pilot programs that monitored these metrics alongside other standard measures (medication adherence, stability of HTN readings & self-monitoring, quality of life, weight/BMI management measures) for risk reduction in HTN-specific care programs.

Our work has a few limitations that warrant further discussion. First, the CCAE database used includes a non-random sample of health plan enrollments; thus, our results may not be nationally representative of general prescription patterns for hypertensive patients in the United States. Second, our treatment patterns analyses were limited to only patterns of drug changes and not dosage changes. Dosage changes may account for some of the treatment patterns observed or may represent additional facets in a patient’s HTN journey that are not captured in our analysis. Third, we have identified that co-medications and co-morbidities are associated with ATL_THREE_ROTO status and the risk of HTN complications within. However, more work needs to done to untangle specific medications and/or comorbidities that are potential drivers of risk within this group of patients. This analysis will be combined with efforts to delineate drivers of ATL_THREE_ROTO status in a future study. Among these “yet to be unraveled” drivers, the complexity of existing treatments is a likely contributor given the excellent correlation between ROTO count with CDS score seen in our study (see **Supplementary Figure 1**); notably, this correlation remained even after removal of HTN drugs for calculation of the CDS score (*data not shown*). As mentioned above, we will evaluate this further in future studies aimed at identifying drivers of ROTO status and the associated risk of HTN complications.

## CONCLUSION

Taken together, our analysis of HTN patients in the United States highlights opportunities for better alignment with current literature-based recommendations (with respect to initial HTN treatment choices) and identified a subset of patients as having elevated HTN risk (based on number of Rx interventions) that may be targeted for clinical/risk management.

## Data Availability

The data used for this manuscript are commercially licensed from CCAE database and not publicly available.

## ACKNOWLEDGEMENTS

We are thankful to members of Geneia LLC’s clinical team, especially, Shelley Riser, Natalie Benner, Hollie Yoder, Ronda Rogers, Mackenzie DeBoer, and Cindy Meck, for their eager, careful insights that guided our treatment pathways analyses and the results presented within this current manuscript.

## DATA AVAILABILITY

**Supplementary Figure 1.**
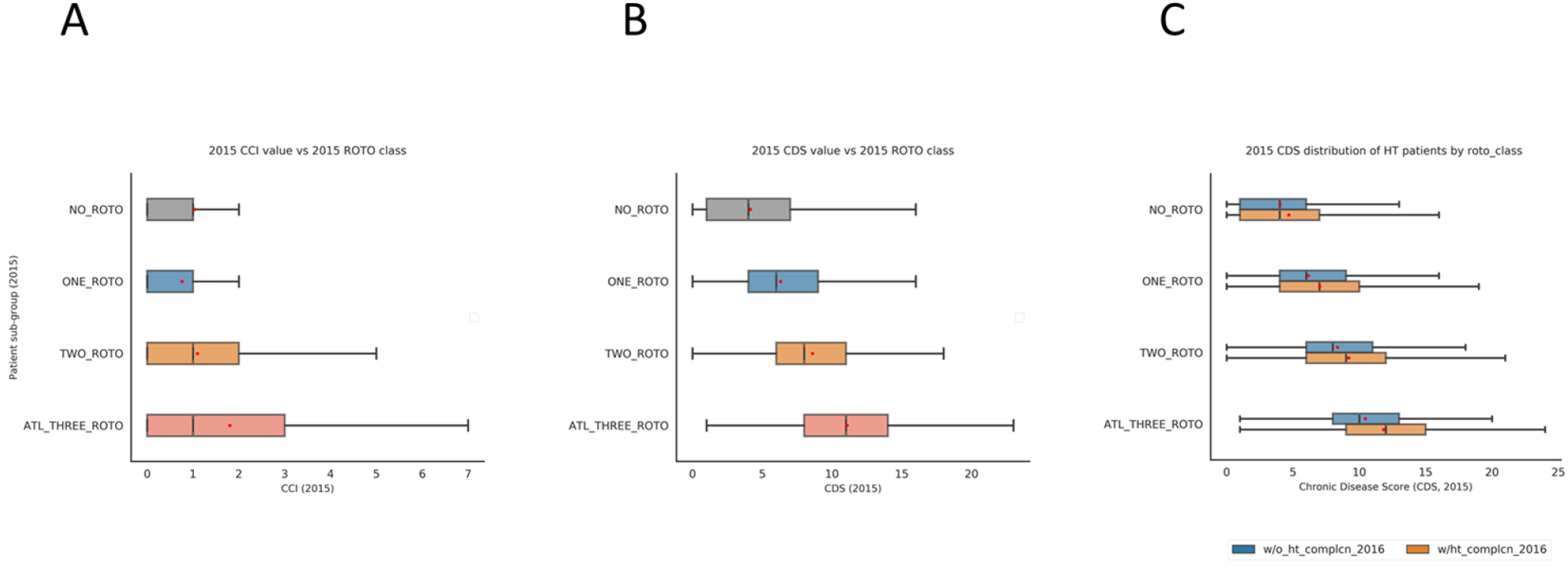
2015 Patient sub-group/ROTO class vs their 2015 comorbidity measures (CCI/CDS distributions) and correlation of 2015 CDS scores with future HTN complications for each ROTO class; CCI = <u>C</u>harlson <u>C</u>omorbidity Index and CDS = <u>C</u>hronic <u>D</u>isease <u>S</u>core. **A**. Boxplot distributions of 2015 CCI values for each 2015 ROTO class. **B**. Boxplot distributions of 2015 CDS values for each 2015 ROTO class. **C**. Distribution of 2015 CDS values by their ROTO class and future HTN complications.

**Supplementary Figure 2.**
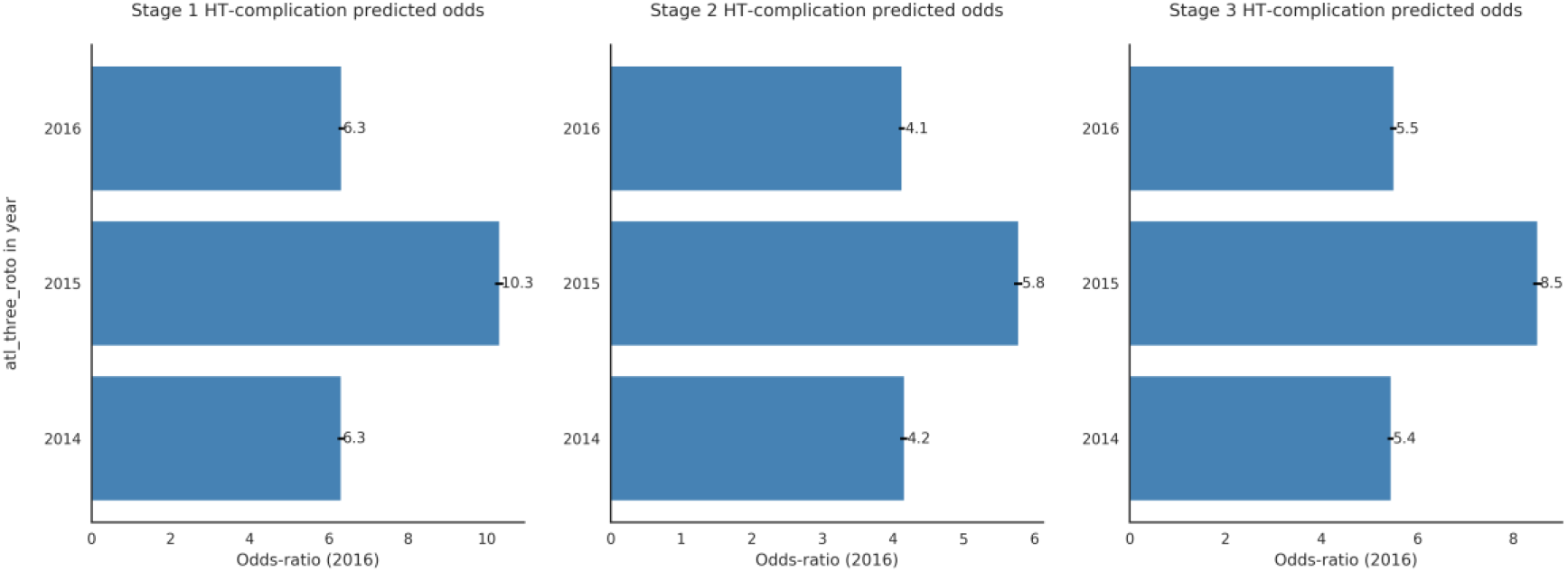
Predicted odds of HTN complications by stage in 2016 and “atl_three_roto” status over current/prior two years (2016, 2015, 2014). “one_roto” group was used as a reference group for relative odds of patients with “atl_three_roto” status. Error bars indicate 95% CI values.

## REFERENCES

1. Benjamin EJ, Muntner P, Alonso A, et al. Heart Disease and Stroke Statistics-2019 Update: A Report From the American Heart Association. Circulation 2019;139(10):e56–e528 doi: 10.1161/CIR.0000000000000659[published Online First: Epub Date]|.

2. Chobanian AV, Bakris GL, Black HR, et al. Seventh report of the Joint National Committee on Prevention, Detection, Evaluation, and Treatment of High Blood Pressure. Hypertension 2003;42(6):1206–52 doi: 10.1161/01.HYP.0000107251.49515.c2[published Online First: Epub Date]|.

3. Lenfant C, Chobanian AV, Jones DW, Roccella EJ, Joint National Committee on the Prevention DE, Treatment of High Blood P. Seventh report of the Joint National Committee on the Prevention, Detection, Evaluation, and Treatment of High Blood Pressure (JNC 7): resetting the hypertension sails. Hypertension 2003;41(6):1178–9 doi: 10.1161/01.HYP.0000075790.33892.AE[published Online First: Epub Date]|.

4. Messerli FH, Williams B, Ritz E. Essential hypertension. Lancet 2007;370(9587):591–603 doi: 10.1016/S0140-6736(07)61299-9[published Online First: Epub Date]|.

5. James PA, Oparil S, Carter BL, et al. 2014 evidence-based guideline for the management of high blood pressure in adults: report from the panel members appointed to the Eighth Joint National Committee (JNC 8). JAMA 2014;311(5):507–20 doi: 10.1001/jama.2013.284427[published Online First: Epub Date]|.

6. Whelton PK, Carey RM, Aronow WS, et al. 2017 ACC/AHA/AAPA/ABC/ACPM/AGS/APhA/ASH/ASPC/NMA/PCNA Guideline for the Prevention, Detection, Evaluation, and Management of High Blood Pressure in Adults: Executive Summary: A Report of the American College of Cardiology/American Heart Association Task Force on Clinical Practice Guidelines. Circulation 2018;138(17):e426–e83 doi: 10.1161/CIR.0000000000000597[published Online First: Epub Date]|.

7. Williams B, Mancia G, Spiering W, et al. 2018 ESC/ESH Guidelines for the management of arterial hypertension. Eur Heart J 2018;39(33):3021–104 doi: 10.1093/eurheartj/ehy339[published Online First: Epub Date]|.

8. Hripcsak G, Ryan PB, Duke JD, et al. Characterizing treatment pathways at scale using the OHDSI network. Proc Natl Acad Sci U S A 2016;113(27):7329–36 doi: 10.1073/pnas.1510502113[published Online First: Epub Date]|.

9. Zhang X, Wang L, Miao S, et al. Analysis of treatment pathways for three chronic diseases using OMOP CDM. J Med Syst 2018;42(12):260 doi: 10.1007/s10916-018-1076-5[published Online First: Epub Date]|.

10. https://www.ibm.com/products/marketscan-research-databases. IBM Market Scan Research Databases. https://www.ibm.com/products/marketscan-research-databases

11. Kulaylat AS, Schaefer EW, Messaris E, Hollenbeak CS. Truven Health Analytics MarketScan Databases for Clinical Research in Colon and Rectal Surgery. Clin Colon Rectal Surg 2019;32(1):54–60 doi: 10.1055/s-0038-1673354[published Online First: Epub Date]|.

12. Halfon P, Eggli Y, Decollogny A, Seker E. Disease identification based on ambulatory drugs dispensation and in-hospital ICD-10 diagnoses: a comparison. BMC Health Serv Res 2013;13:453 doi: 10.1186/1472-6963-13-453[published Online First: Epub Date]|.

13. Quan H, Sundararajan V, Halfon P, et al. Coding algorithms for defining comorbidities in ICD-9-CM and ICD-10 administrative data. Med Care 2005;43(11):1130–9 doi: 10.1097/01.mlr.0000182534.19832.83[published Online First: Epub Date]|.

14. McGregor JC, Kim PW, Perencevich EN, et al. Utility of the Chronic Disease Score and Charlson Comorbidity Index as comorbidity measures for use in epidemiologic studies of antibiotic-resistant organisms. Am J Epidemiol 2005;161(5):483–93 doi: 10.1093/aje/kwi068[published Online First: Epub Date]|.

15. Von Korff M, Wagner EH, Saunders K. A chronic disease score from automated pharmacy data. J Clin Epidemiol 1992;45(2):197–203 doi: 10.1016/0895-4356(92)90016-g[published Online First: Epub Date]|.

16. McCammon J et al. Use of machine learning to predict hypertension-related complication outcomes of varying severity manuscript in preparation 2020

17. Robles NR, Cerezo I, Hernandez-Gallego R. Renin-angiotensin system blocking drugs. J Cardiovasc Pharmacol Ther 2014;19(1):14–33 doi: 10.1177/1074248413501018[published Online First: Epub Date]|.

18. Suchard MA, Schuemie MJ, Krumholz HM, et al. Comprehensive comparative effectiveness and safety of first-line antihypertensive drug classes: a systematic, multinational, large-scale analysis. Lancet 2019;394(10211):1816–26 doi: 10.1016/S0140-6736(19)32317-7[published Online First: Epub Date]|.

19. Wright JM, Musini VM, Gill R. First-line drugs for hypertension. Cochrane Database Syst Rev 2018;4:CD001841 doi: 10.1002/14651858.CD001841.pub3[published Online First: Epub Date]|.

20. Wang G, Chen Y, Li L, Tang W, Wright JM. First-line renin-angiotensin system inhibitors vs. other first-line antihypertensive drug classes in hypertensive patients with type 2 diabetes mellitus. J Hum Hypertens 2018;32(7):494–506 doi: 10.1038/s41371-018-0066-x[published Online First: Epub Date]|.

21. Zhang X, Zhao Q. Association of Thiazide-Type Diuretics With Glycemic Changes in Hypertensive Patients: A Systematic Review and Meta-Analysis of Randomized Controlled Clinical Trials. J Clin Hypertens (Greenwich) 2016;18(4):342–51 doi: 10.1111/jch.12679[published Online First: Epub Date]|.

